# The COVID-19 Spread in the State of Assam, India

**DOI:** 10.1101/2020.09.18.20197095

**Authors:** Hemanta K. Baruah

## Abstract

We have studied the current COVID-19 spread situation in Assam, a State of India. We have found that currently the spread pattern is indeed exponential and that it is not going to show a reducing trend soon. As a result, it is not possible yet to forecast about the time of peaking of the epidemic in Assam. It can be said that the COVID-19 situation in this Indian State is very alarming even after five and a half months of the start of the epidemic in the State. It may so happen that in Assam the spread would continue to grow exponentially even after the situation changes in India as a whole.

## Introduction

The commonly used mathematical models used for forecasting the spread of an epidemic are the Susceptible-Infectious-Recovered (SIR) model [1, 2, 3] and its modifications such as the Susceptible-Exposed-Infectious-Recovered (SEIR) model, the Susceptible-Infectious-Recovered-Dead (SIRD) model, and the Susceptible-Infectious-Susceptible (SIS) model. Due to certain necessary assumptions made in these models, it is debatable whether such data dependent models are everywhere applicable in the COVID-19 matters [4]. We shall in this article study the current COVID-19 status as far as the spread is concerned in the State of Assam, India, without using any standard epidemiological model.

We shall first describe some associated matters related to Assam so that the readers get an idea about its demographic details. Assam is a State of India, located in the North-East. As per the data of Census of India, 2011, the population of Assam was 31.2 million. As per Unique Identification India data, updated on 31 May, 2020, the projected population of Assam is 35.6 million. The total area of the State is 78,438 sq km, with population density 397 per sq km.

Regarding COVID-19 spread, the daily data of the cumulative total number in India can be easily found from Worldometers.info [5]. However this source shows the concerned data with reference to Assam not from the beginning but from 16 August onwards. To collect the concerned data of Assam from the beginning, we had to take resort to the following. From March 31, 2020, the date on which the first COVID-19 patient was detected in Assam, to August 19, 2020, the daily data are available in the COVID-19 Pandemic in Assam portal [6]. The data available in this portal were not updated after August 19. Therefore to get the data thereafter, we had to rely on the data made available online by the Assam COVID-19 Dashboard, Government of Assam [7]. However, in this portal data for every current day only are available. So as to get the earlier data, one had to keep track of the updates everyday, which we have done, and we have the concerned data till the present day. As has been mentioned earlier, the daily data are available in the Worldometers.info [4] from August 16 anyway.

It was mentioned in [4] that unless the data regarding recovered cases and death are reliable enough, the epidemiological models such as SIR, SIS and SIRD cannot be blamed. The population exposed to the disease may not have economic homogeneity, and in that situation the SEIR model similarly cannot be blamed for unacceptable results.

It is known that patients with comorbidities should take all necessary precautions to avoid getting infected with the SARS CoV-2 as they have the worst prognosis (see for example [8]). In Assam, it has been reported that the number of deaths due to SARS CoV-2 does not include death of corona patients with comorbidity after recovery as death due to the corona virus disease. Indeed this might actually have been the followed norms elsewhere also the world over. This in turn shall make the projections using the standard epidemiological models faulty.

Further, in a region with high population density, how well the people are maintaining social distance is an important point. Indeed, due to parameters associated with poverty, maintaining social distance cannot be homogeneous anyway. Finally, although at the initial stage medical facilities were more readily available to the corona patients, currently due to the high spread of the disease, medical facilities are getting reduced. This might end up affecting the recoveries.

We shall in this article analyze the data of the spread of the virus in Assam assuming that the spread pattern is a function of time only. We shall not study the situation using any standard epidemiological model. We shall first see whether the cumulative total number of cases is following the exponential pattern at least approximately. Thereafter we shall see if forecasts can be made regarding the total number of cases in Assam.

## The Data

We have already mentioned that the data regarding the total number of COVID-19 cases are readily available on the web by the name COVID-19 Pandemic in Assam, from March 31 to August 19. We are now going to display the data from August 20 onwards collected on a daily basis from the portal Assam COVID-19 Dashboard, Government of Assam. These data tally with the Worldometers.info data displayed from 16 August onwards. We are interested to display the data from August 20 to September 14 because the Assam Government Portal gets updated every single day, and on any day the data for that day only can be easily seen. We have actually analyzed the data from August 22 to September 10. The data for the four days thereafter would be of help to compare the forecasts with the actual values.

**Table 1:** Total Number of Cases from 20 August to 14 September.

| Dates | Cases | Dates | Cases |
| --- | --- | --- | --- |
| 20-Aug | 86052 | 2-Sep | 115279 |
| 21-Aug | 87908 | 3-Sep | 118333 |
| 22-Aug | 89468 | 4-Sep | 121224 |
| 23-Aug | 90740 | 5-Sep | 123922 |
| 24-Aug | 92619 | 6-Sep | 125459 |
| 25-Aug | 94592 | 7-Sep | 128244 |
| 26-Aug | 96771 | 8-Sep | 130823 |
| 27-Aug | 98807 | 9-Sep | 133066 |
| 28-Aug | 101367 | 10-Sep | 135805 |
| 29-Aug | 103794 | 11-Sep | 138339 |
| 30-Aug | 105774 | 12-Sep | 140471 |
| 31-Aug | 109040 | 13-Sep | 141763 |
| 1-Sep | 111724 | 14-Sep | 144166 |

**Fig. 1:**
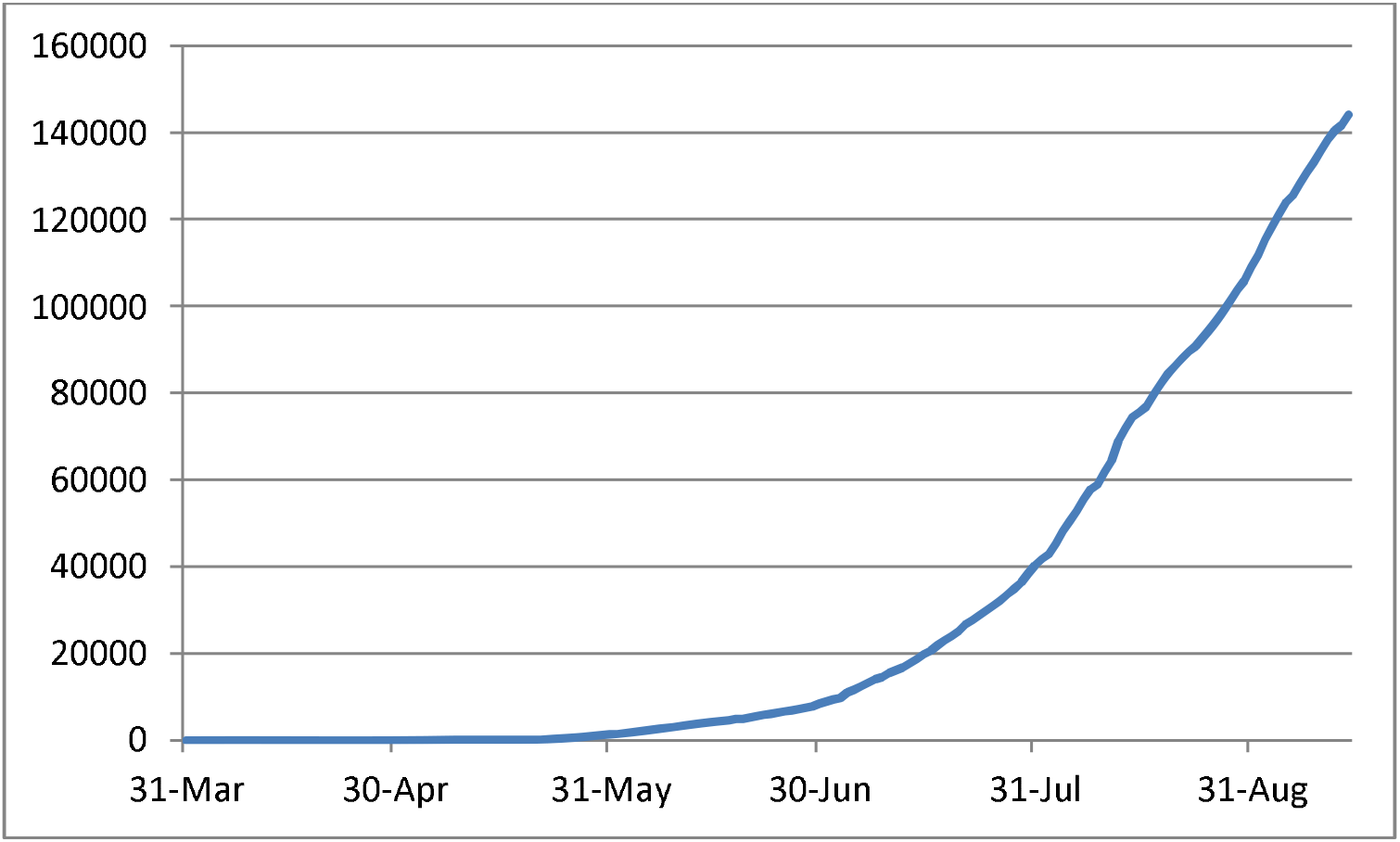
Plot of the COVID-19 Spread Data from 31 March to 14 September.

From the figure above, we can surmise that the growth pattern is perhaps exponential.

## The Analysis

We first hypothesize that the cumulative total number of cases in Assam is following an exponential pattern. To test the acceptability of this hypothesis we shall proceed as follows. We shall test whether the function

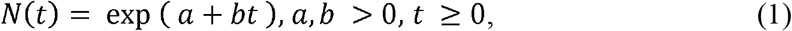

fits the data of spread in Assam approximately, where *N(t)* stands for the cumulative total number of cases on day *t*, and *a* and *b* are parameters to be estimated. Let *Z=logeN(t)* Now if the estimated linear equation defining *Z(t)* can be found to be statistically acceptable, that would mean that there should be no reason to reject our hypothesis stated above.

Before testing for acceptability of the hypothesis, we would like to have a look at whether the values of Δ *Z(t)*, the first order differences of *Z(t)*, are showing a reducing trend. Let us write Δ *Z(t)*= α + β *t*. If the estimate of *β* comes out to be negative, then we would test the acceptability of the fitted regression equation of Δ*Z(t)* on *t*. If the equation is found to be statistically acceptable, then it can be used for forecasting. In that kind of a case, testing for validity of equation (1) would not be necessary. However, if the equation is found to be rejectable, or if the estimated value of the parameter *β* is found to be positive, then we would go for testing the validity of equation (1) statistically so that it can be used for forecasting.

Indeed, although in the applications of the epidemiological models it is assumed that in the second stage of the spread the pattern is exponential, it can only be approximately exponential and not absolutely exponential because that would mean that the epidemic would never end. Now in the approximately exponential stage, the parameter *b* in (1) above cannot actually remain constant for a long period, it has to have a decreasing trend.

At this point, we would like to mention that in the case of the World outside China, it was seen in that during the period of the study, the average values of Δ*Z(t)* had followed the following pattern:

A. 0.13705 from March 21 to March 28,
B. 0.09287 from March 29 to April 4,
C. 0.05916 from April 5 to April 11,
D. 0.04078 from April 12 to April 18, and
E. 0.03181 from April 19 to April 25.

What we mean is that the parameter *b* in (1) was seen to have a decreasing trend. It was mentioned in [4] that the average value of Δ*Z(t)* in India during the 14 days from May 11 to May 24 was 0.051716. It was seen further that during the 7 days from May 25 to May 31, the average value of Δ*Z(t)* was 0.045584. The average value of Δ*Z(t)* in India [4] from June 1 to June 7, 2020, was 0.04063. From June 8 to June 10, the average came down to 0.03635. Therefore the reducing trend of Δ*Z(t)* in the approximately exponential stage of the outbreak was actually observed.

In the following table, we are going to show the values of Δ*Z(t)* for Assam for the duration of 20 days from August 22 to September 10.

**Table 2:**
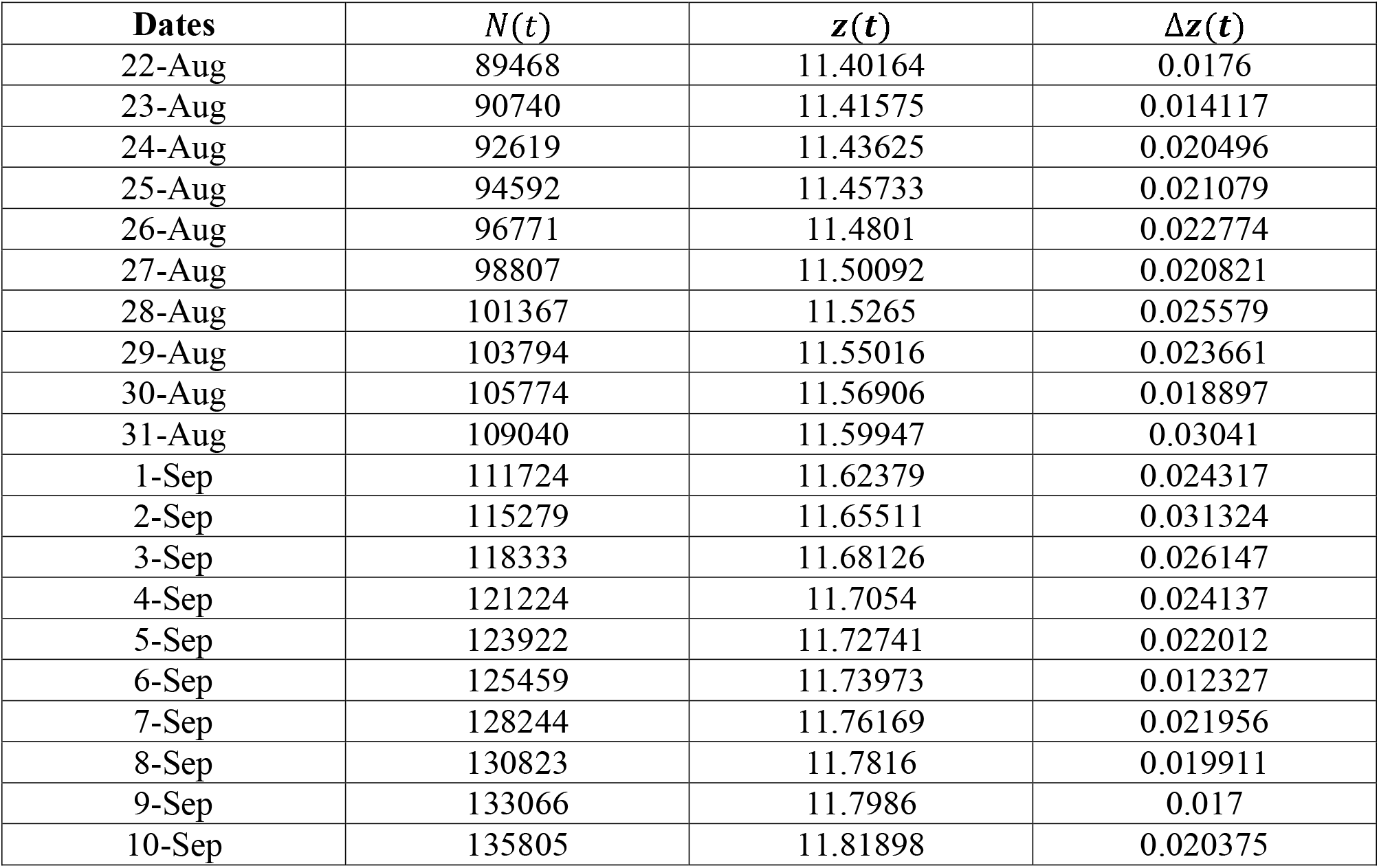
Values of ΔZ(t) from 22 August to 10 September.

From Fig. 2 it can be surmised that perhaps the values of Δ*Z(t)* are not showing a reducing trend. Indeed, the regression equation of Δ*Z(t)* on *t* was found to be

**Fig. 2:**
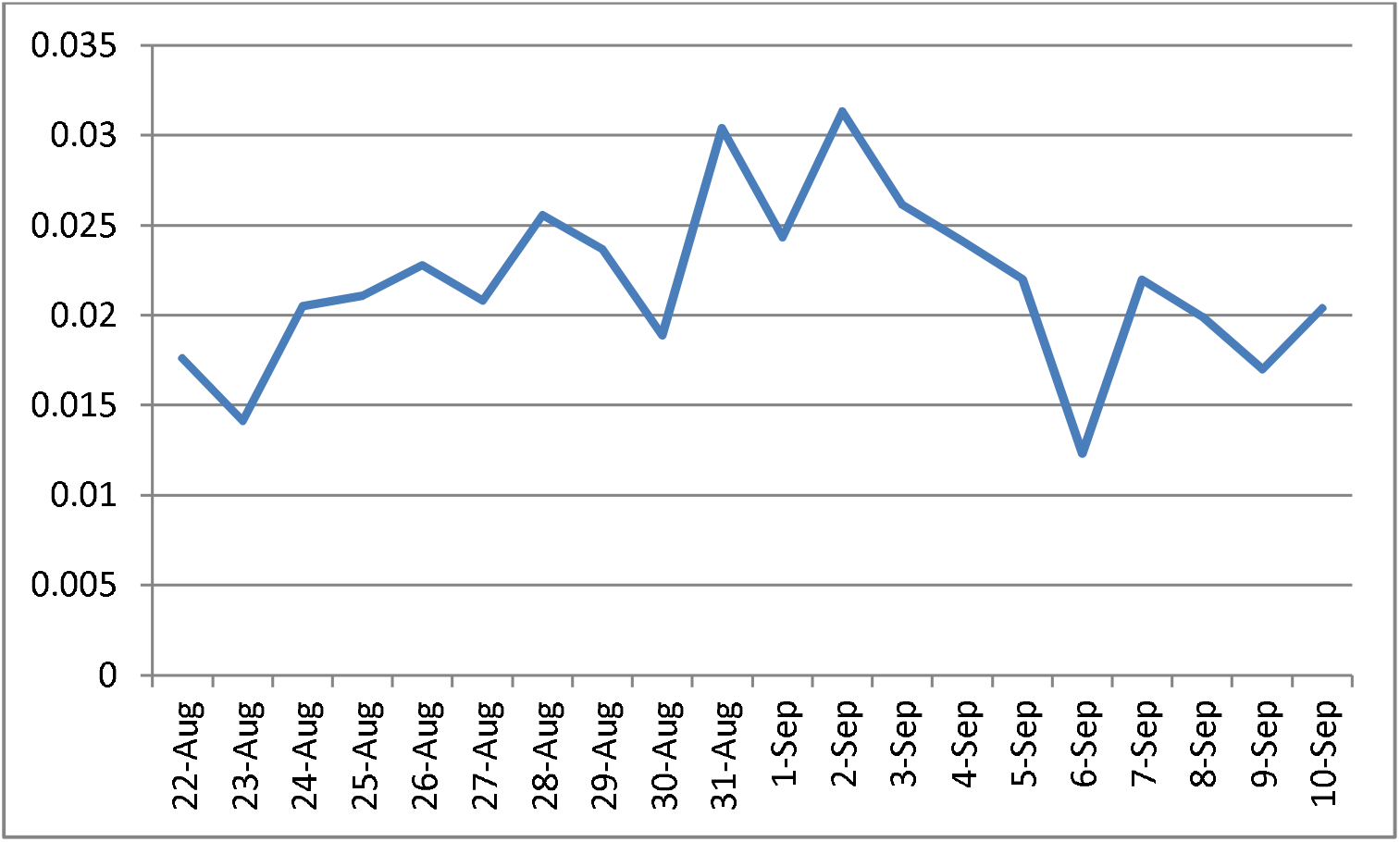
Graphical Representation of ΔZ(t) from 22 August to 10 September.

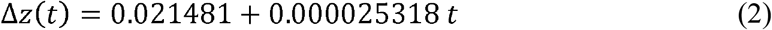

As the estimate of *β* has been found to be positive (= 0.000025318), we can say that during the period from August 22 to September 10 the variable Δ*Z(t)* has actually shown an increasing trend. We would have gone for a statistical test of validity of this equation had the estimate of *β* been negative. But as surmised, the estimate has come out to be positive, and therefore testing for validity of this equation statistically is of not much use.

What we have found is very alarming. As was discussed earlier, Δ*Z(t)* should show a decreasing trend, which asserts that the cumulative total number of cases is indeed following an almost exponential pattern of growth and also that the pattern would in course of time change towards an almost logarithmic pattern as happens in the cases of epidemics. However in Assam during the period of those 20 days, the cumulative total number of cases has not reflected a tendency towards improvement. The estimate of *β* being positive points to this kind of a conclusion only. We shall now have to see whether the spread pattern is actually of the exponential type or not. It may be that the pattern is just highly nonlinear and is not nearly exponential yet.

We shall therefore move forward to examining whether equation (1) has validity during those 20 days. We shall therefore check whether the regression equation of *Z(t)* on t, *Z(t)* =a + *bt* is statistically valid. The regression equation fitted using the method of least squares was found to be for t = 1,2,…,

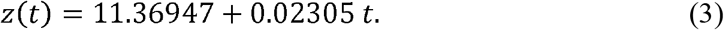

Using the Student’s *t* test, with

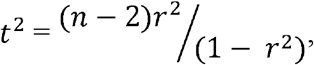

where *n*= 20, we have found that the calculated value of *t* comes out as equal to 78.089 which is far greater than the two sided theoretical value of *t* (= 2.101), at 5% probability level of significance for 18 degrees of freedom. We have indeed done the statistical test of the null hypothesis H0 : *ρ*= 0 against the two sided alternative hypothesis H1ρ ≠0 where is the population correlation coefficient between the variables Δ*Z(t)* and *t*. Therefore we conclude that during this period the null hypothesis is to be rejected, and that there is a significant linear relationship between *Z(t)* and t. Using equation (3) we have calculated the expected values of *N(t)*. The diagrammatic representation has been shown in Fig. 3. In the figure, the first series represents the observed values and the second series represents the expected values. From Fig. 3, it can be seen that the expected values have started to slightly overestimate from 8 September onwards. So if we make forecasts with the help of the regression equation (3), we may end up overestimating the cumulative total number of cases. We have earlier tried to forecast the total number of cases ([4], [9]) using the average value of Δ*Z(t)* successfully. As Δ*Z(t)* in the current case has not shown a decreasing trend, we shall not go for forecasting with reference to Δ*Z(t)* because in that way also we shall end up making overestimating forecasts only. In Table-3 we have shown the forecasts for 20 days starting from 11 September using equation (3).

**Table 3:** Forecasts Based on the Regression Equation.

| Dates | Forecasts |
| --- | --- |
| 11-Sept | 140589 |
| 12-Sept | 143868 |
| 13-Sept | 147223 |
| 14-Sept | 150657 |
| 15-Sept | 154170 |
| 16-Sept | 157766 |
| 17-Sept | 161445 |
| 18-Sept | 165210 |
| 19-Sept | 169063 |
| 20-Sept | 173005 |
| 21-Sept | 177041 |
| 22-Sept | 181169 |
| 23-Sept | 185394 |
| 24-Sept | 189718 |
| 25-Sept | 194143 |
| 26-Sept | 198670 |
| 27-Sept | 203304 |
| 28-Sept | 208045 |
| 29-Sept | 212897 |
| 30-Sept | 217862 |

**Fig. 3:**
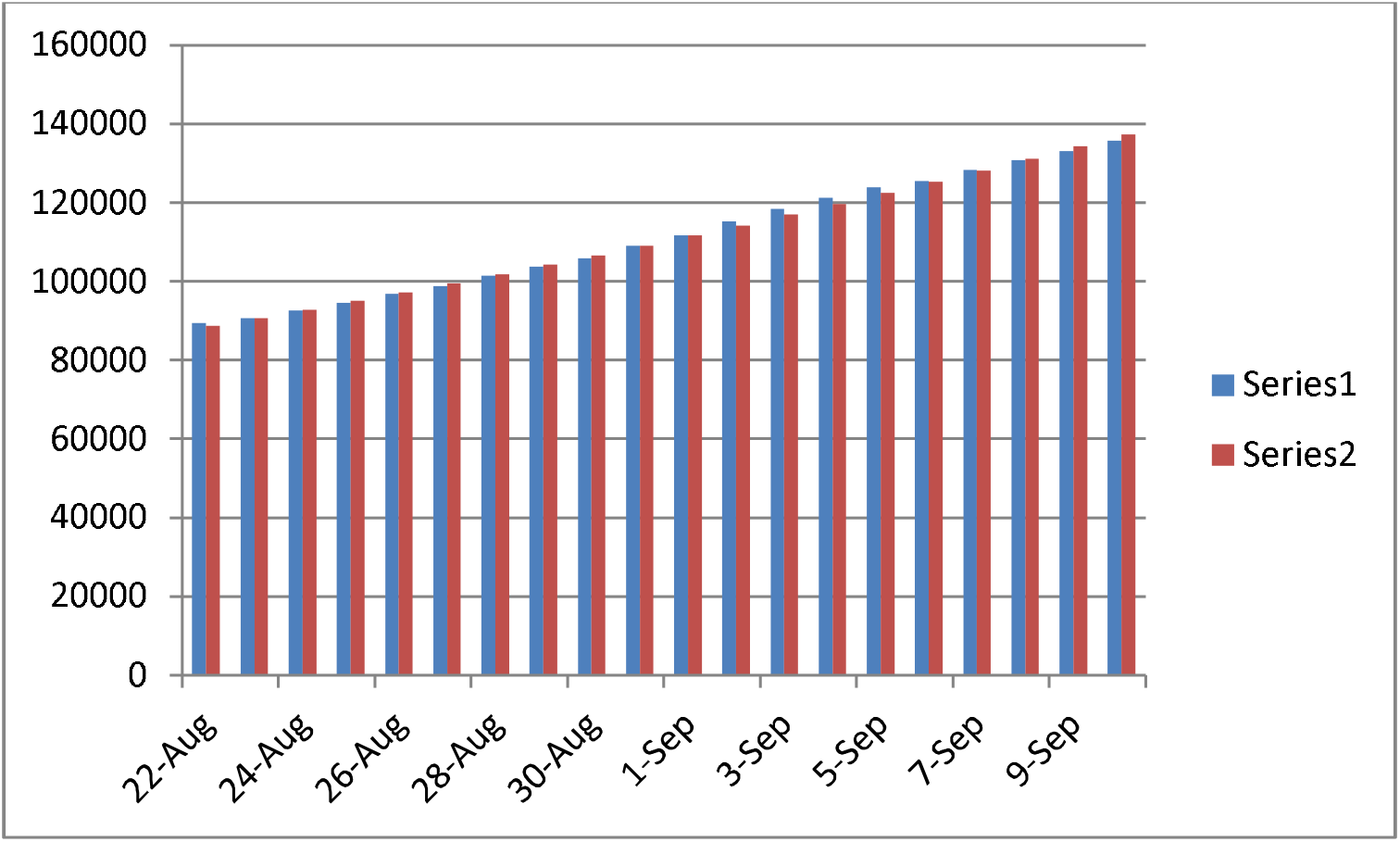
Observed and Expected Values of N(t) from August 22 to September 10.

It may be noted that in Italy, the epidemic appeared on February 15, and after just two and a half months by the end of April the process of curve flattening had already started [10]. In India as a whole also the epidemic appeared on February 15, and it is expected that the process of curve flattening is about to start [4]. However, though in Assam the epidemic had started on March 31, even now it is exponentially growing without showing any signs of retardation. It can be said that the COVID-19 situations in India and in one of its States, Assam, are totally different. In other words, the spread situations in India as a whole and in a small State of India are not really comparable. It may so happen that in Assam the spread would continue to grow exponentially even after the situation changes in India as a whole.

## Conclusions

We have found that the COVID-19 situation in Assam, India, is very grim. The current pattern followed by the cumulative total number of cases *N(t)* is exponential of the type exp(a+ *bt*). But the parameter *b* is not showing a reducing trend yet, which is shown in all epidemiological cases. Until the parameter *b* starts showing a reducing trend, nothing can be said when exactly the epidemic would start peaking. In India as a whole, this reducing trend in *b* was found to have started in the first half of May, and the spread in the country is still on the increase. In Assam, as the reducing trend is yet to be seen, to what kind of a dangerous stage it would continue to spread cannot be mathematically foreseen even now. In Italy for example, after two and a half months from the start, the process of retardation had started. In Assam, even after five and a half months from the start, the epidemic is still in an accelerating and unpredictable stage.

## Data Availability

COVID-19 Pandemic in Assam portal, Worldometers.info, Assam COVID-19 Dashboard, Government of Assam.

